# Real-World Benchmarking and Validation of Foundation Model Transformers for Endometrial Cancer Subtyping from Histopathology

**DOI:** 10.1101/2025.10.10.25337691

**Authors:** Vincent M. Wagner, Casey M. Cosgrove, Stephanie J. Chen, Daniel T. Griffin, Megan I. Samuelson, Michael J. Goodheart, Jesus Gonzalez-Bosquet

## Abstract

To evaluate whether open-source histopathology foundation model pipelines, paired with attention-based multiple instance learning (MIL), can accurately classify molecular subtypes of endometrial cancer (EC) from whole-slide images (WSIs) and maintain performance in a real-world, independent cohort.

**Methods:** We assembled a public discovery cohort of 815 patients (1,195 WSIs) from The Cancer Genome Atlas and Clinical Proteomic Tumor Analysis Consortium, and an independent external cohort of 720 patients (1,357 WSIs) with molecular subtyping determined by mismatch repair immunohistochemistry plus TP53 and POLE sequencing. Four ImageNet-pretrained convolutional neural networks (CNNs) and six open-source foundation encoders using two MIL aggregation strategies (TransMIL and CLAM) were benchmarked within the STAMP pipeline. Models were trained with five-fold cross-validation and evaluated on an independent cohort. Macro–area under the receiver operating characteristic curve (AUC) was the primary outcome.

**Results:** In cross-validation, foundation models outperformed CNNs (macro-AUC 0.799-0.860 vs 0.715-0.829). The best configuration (Virchow2 with CLAM) achieved macro-AUC 0.860 (95%CI, 0.839-0.880), macro-F1 score 0.607, and balanced accuracy 0.647. External validation showed substantial degradation for CNNs, while foundation models retained higher discrimination (macro-AUC 0.667-0.780). UNI2 with CLAM had the highest external macro-AUC (0.780), and Virchow2 with CLAM had the best balanced accuracy (0.525). Subtype-level AUCs for UNI2 with CLAM were highest for p53abn (0.851).

**Conclusions:** Open-source foundation model pipelines with attention-based MIL can deliver accurate and generalizable molecular subtyping of EC directly from WSIs. These models outperform CNNs in real-world validation, supporting their potential as scalable, cost-effective tools to guide precision oncology and triage confirmatory molecular testing.

## Introduction

Endometrial cancer (EC) is the most frequently diagnosed gynecologic cancer in the United States and, unlike many other solid tumors, its incidence and mortality are on the rise^1, 2^. This trend represents a growing clinical and societal burden. While histology and tumor stage has traditionally guided treatment decisions^3^, histology has high inter-observer disagreements and both offer limited utility in the context of personalized or molecularly targeted therapies^4^. In 2013, The Cancer Genome Atlas (TCGA) introduced a landmark classification system for EC, identifying four prognostically different molecular subtypes: polymerase ε (POLE) ultra-mutated, high microsatellite instability hypermutated (MSI-high), copy-number low (CNV-L), and copy-number high (CNV-H)^5^. The prognostic distinction between the subtypes have been consistently demonstrated in multiple patient cohorts using more clinically accessible testing strategies^6–8^.

To facilitate clinical adoption, surrogate assays combining mismatch repair (MMR) and p53 protein immunohistochemistry (IHC) for MSI-high and CNV-H, respectively, along with targeted *POLE* sequencing have been developed with analogous molecular subtypes: MMR-deficient (dMMR), p53 aberrant (p53abn), *POLE* pathogenic mutation (POLE) and no specific molecular profile (NSMP)^7, 9^. This surrogate-based classification has been incorporated into treatment guidelines, staging criteria and increasingly being used in practice as well as in clinical trials^10–12^. However, these surrogate algorithms do not correlate perfectly with the TCGA methods and routine complete molecular classification has not yet been universally implemented, limited by cost, complexity and turnaround time^13, 14^.

Hematoxylin and eosin (H&E)-stained slides, produced for every patient at diagnosis, contain histologic features that reflect these molecular subtypes. While expert pathologists can identify some morphologic correlates, deep learning (DL) and artificial intelligence (AI) methods can uncover subtle, more complex morphologic and spatial signatures that are beyond human perception^15^. Studies have demonstrated the potential of AI to extract prognostic and genomic information from H&E whole-slide images (WSIs) across a range of cancer types^16^.

Several groups have explored AI-based molecular classification in EC using many different AI architectures, DL techniques and datasets highlighting the promise of AI^17–22^. However, these studies are often constrained by proprietary model architectures, lack of external validation, or reliance on non-public datasets. Furthermore, none of these have systematically compared different architectures, encoders and aggregators to benchmark the state-of-the-art models.

State-of-the-art histopathology foundation models offer a powerful alternative to fully in-house trained models. These general purpose histology feature extractors are trained on millions of histology tiles using self-supervised learning and can be repurposed for downstream tasks without full retraining^23^. These encoders require the WSIs to be split into tiles for embedding, which then need to be aggregated for whole slide prediction.

To achieve this under weak supervision (i.e. without tile-level labels), multiple-instance-learning (MIL) is used. Among the leading open-source MIL strategies are TransMIL, a transformer-based aggregator using inter-tile self-attention^24^, and CLAM, which applies class-specific attention pooling and instance-level supervision^25^. Both approaches have demonstrated high performance in histopathology tasks while offering architectural flexibility and interpretability. The recently introduced STAMP (Solid-Tumour Associative Modelling in Pathology) framework offers an open-source pipeline that combines tiling, feature extraction using foundation models and MIL-based prediction^26^. This enables rigorous benchmarking of AI pipelines using community-shared encoders and datasets.

The objective of this study was to systematically benchmark six foundation encoders, paired with two MIL aggregation methods (TransMIL and CLAM) against standard CNNs, using the STAMP pipeline, Figure 1. We trained and evaluated these models on a public discovery cohort using TCGA and Clinical and Proteomic Tumor Analysis Consortium (CPTAC) and tested generalizability on an independent, real-world cohort of over 700 patients. We hypothesized that foundation encoder-based pipelines could deliver accurate and cost-effective molecular classification directly from H&E slides, potentially enabling scalable, point-of-care precision oncology for patients with endometrial cancer.

**Figure 1:**
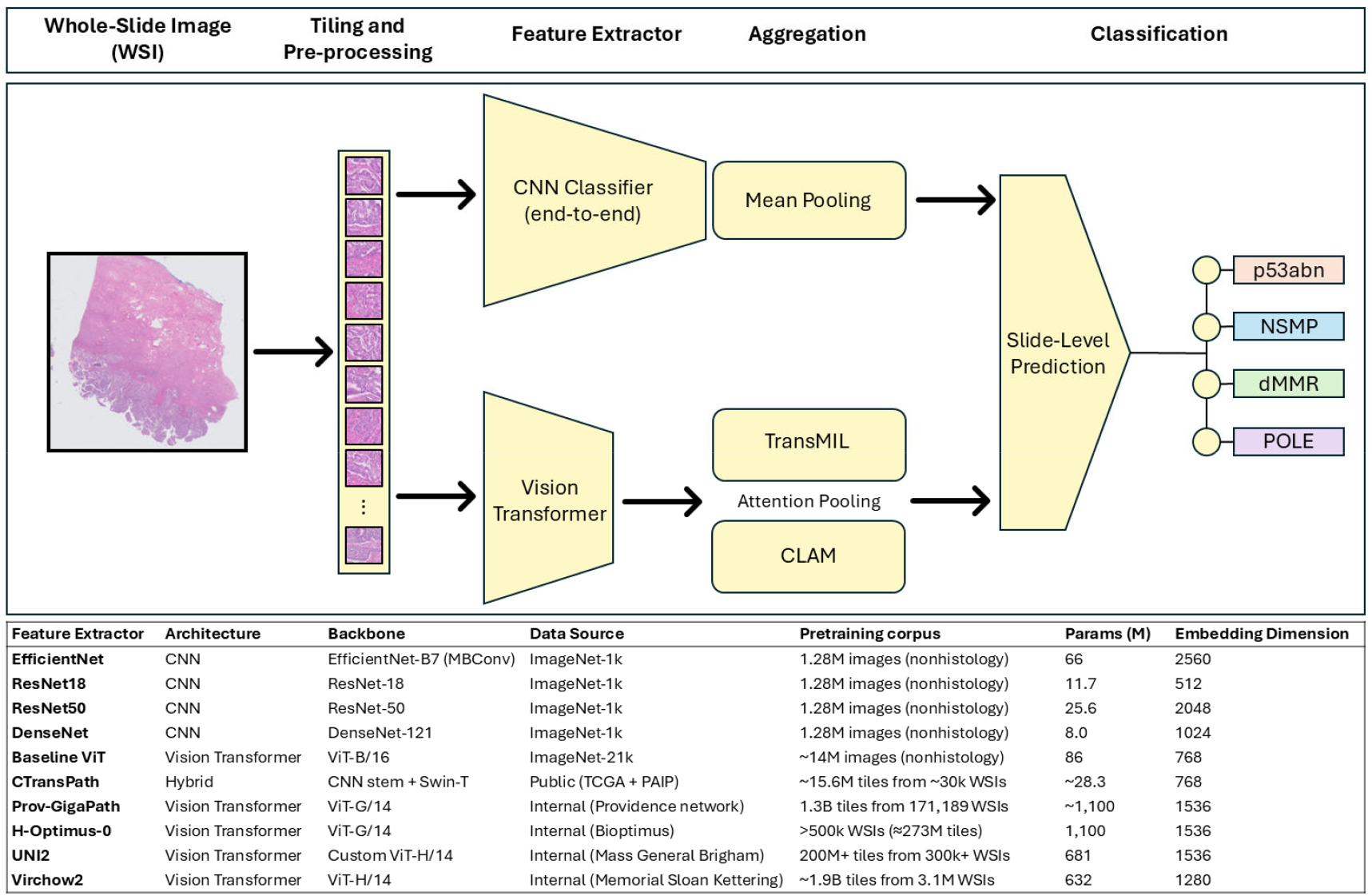
Schematic and Foundation Feature Extractors. Overview of the computational pathology workflow for molecular subtype prediction from whole-slide image including the convolutional neural network pipeline and vision transformer pipeline with attention pooling using TransMIL or CLAM. The bottom includes a list of the feature extractors used along with information about each including architecture, backbone, training data, parameter count (M) and embedding dimension. Abbreviations: CNN, convolutional neural network; ViT, vision transformer; WSI, whole-slide image; MIL, multiple instance learning; CLAM, clustering-constrained attention multiple instance learning; TCGA, The Cancer Genome Atlas; PAIP, Pathology AI Platform.

## Methods

### Data sources and cohort assembly

#### Public discovery cohort

WSIs and matched molecular data were downloaded from TCGA and CPTAC; TCGA-UCEC (N=523), TCGA-UCS (N=52), and CPTAC-UCEC (N=240) projects. All images and data from the TCGA and CPTAC are publicly available at https://portal.gdc.cancer.gov and https://www.cancerimagingarchive.net. After compiling, 815 unique patients were identified. These individuals contributed 1195 diagnostic H&E WSIs (median 1 slide per patient, IQR 1-2). Molecular sub-types, patient and slide-level distribution in Table 1. Slides were randomly split (stratified by subtype) into training (70%), validation (15%) and internal test (15%) sets for cross-validation experiments.

**Table 1:**
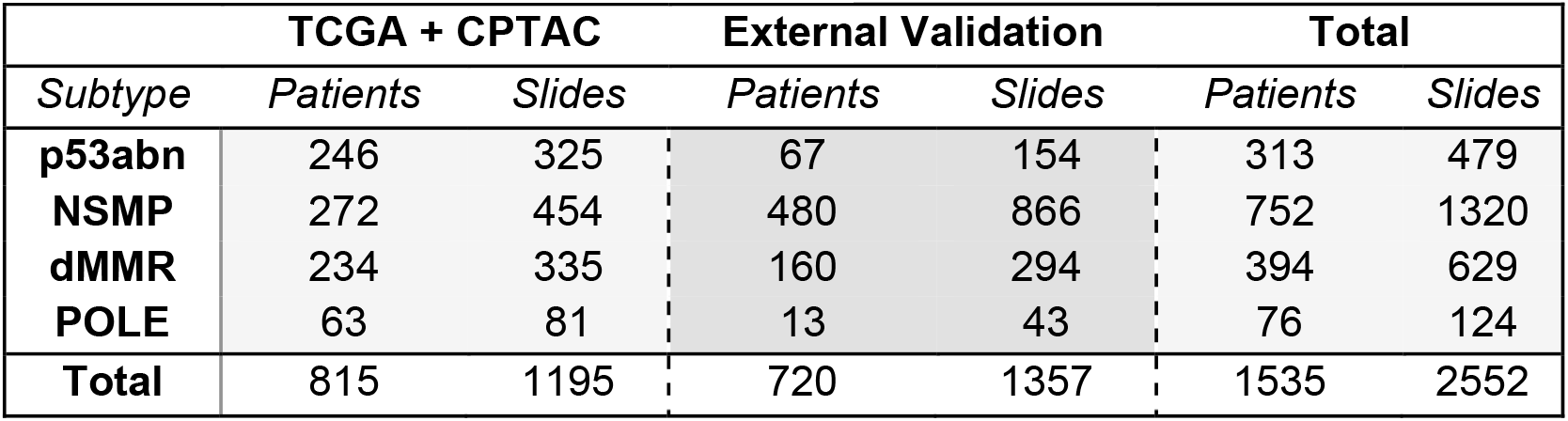
Training and Validation Cohorts by Subtype.

#### Independent external cohort

An independent cohort was assembled under IRB approval including 720 patients with endometrial cancer between 2014-2017^27^, yielding 1357 WSIs with confirmed molecular labels, Table 1. Molecular subtype was determined using IHC for MMR, MSI testing using consensus microsatellite markers, and NGS for TP53 and POLE^27^ (only known pathogenic mutations were labeled positive^28^). No slides or patients from this cohort were used during model development; they were reserved exclusively for held-out deployment testing to evaluate generalization performance of the models from cross-validation.

#### Preprocessing

All WSIs were processed with a modified HIA^29^ and STAMP pipeline^26^. Slides were not manually annotated or segmented for tumor or region of interest. WSIs were tessellated into patches (256 x 256 microns of tissue). WSI were filtered with two quality checks: background filter to exclude background patches (brightness filter) and texture filter using canny edge mask effectively removing out-of-focus regions.

#### CNN Pipeline

We evaluated four CNN architectures with ImageNet^30^-pretrained weights (EfficientNet-B7^31^, ResNet-18^32^, ResNet-50^32^, and DenseNet^33^) for tile-level classification. Architectures were selected to span a range of depth, connectivity, and computational complexity. A portion of layers was frozen, and the remainder fine-tuned with optimization under five-fold cross-validation. Slide-level predictions were generated by mean-pooling tile outputs. Top-scoring tiles and slides were retained for interpretability analysis. Complete architecture specifications, hyperparameters, and training configurations in supplementary appendix 2.

#### Transformer and Multiple Instance Learning (MIL) Pipeline

For the transformer pipeline, tiles were embedded using frozen (weights kept constant) encoders/feature extractors (ViT-B/16^34^, CTransPath^35^, Prov-GigaPath^36^, H-Optimus-0^37^, UNI-2^38^, Virchow2^39^) which were downloaded through https://huggingface.co/. Embeddings were aggregated into slide-level predictions using either a transformer-based MIL aggregator (TransMIL) or an attention-based pooling model with instance-level supervision (CLAM-MB). Both pipelines were trained with five-fold cross-validation using fixed bag sizes (number of tiles) and frozen encoders with only the aggregator trainable. Full model configurations and hyperparameters in supplementary appendix 2.

#### Statistics

Performance was summarized for each cross-validation fold and for the five external validation folds of the independent test set. The primary outcome metric was the macro-ROC-AUC (macro-AUC), defined as the average of the per-subtype ROC-AUCs, giving equal weight to each molecular subtype regardless of prevalence. As complementary metrics, we also calculated the macro-F1 score (the average F1 score across subtypes, balancing precision and recall) and balanced accuracy (the mean recall across subtypes), both of which account for class imbalance by ensuring minority subtypes are weighted equally. In addition, per-subtype ROC-AUCs were evaluated as secondary outcomes to provide subtype-specific discrimination performance. We report means with 95% confidence interval (95%CI). To assess whether observed differences in macro-AUC were statistically significant, we performed two-sided paired t-tests on the fold-level metric vectors. Normality of fold-wise deltas was verified with the Shapiro– Wilk test (P > 0.10 for all contrasts). Resulting p-values were adjusted for multiple testing with Benjamini-Hochberg false-discovery-rate (FDR) (α = 0.05). Effect sizes are presented as ΔAUC with 95%CI.

#### Interpretability analysis

To ensure biological plausibility and clinical relevance, we included interpretability analyses: qualitative review of attention maps/tiles and quantitative evaluation of cell composition and nuclear morphometrics. This dual approach verifies models focus on expected histology and provides objective, reproducible measures. We selected the 10 highest predicted WSI from each subtype and then identified the 8 highest scoring tiles from each WSI using attention heatmaps for the correctly-predicted subtype, yielding 80 tiles per subtype for interpretability analysis. Tiles were analyzed by trained pathologists. Nuclei were then segmented with TIAToolbox^40^ using the HoVer-Net^41^ with PanNuke^42^ schema and nuclear morphometric features were extracted from neoplastic epithelial cells to quantify size variability, pleomorphism, eccentricity and circularity.

Differences among subtypes were evaluated with the Kruskal–Wallis test, followed by Dunn’s post-hoc comparisons with Holm correction. Full feature definitions, computational settings, and visualization methods in supplementary appendix 2.

## Results

### Cross-validation

During five-fold cross-validation every architecture achieved a macro-AUC above 0.71, confirming discriminative learning, Figure 2 (light bars), Table S1. Among the CNNs, EfficientNet was clearly the weakest with macro-AUC 0.715 (95%CI 0.675-0.754), macro-F1 score 0.404 (95%CI 0.353-0.455), balanced accuracy 0.420 (95%CI 0.372-0.468), while ResNet-18, ResNet-50 and DenseNet clustered at macro-AUC 0.81-0.83 (95%CI width≈0.05) with macro-F1≈0.58 and balanced accuracy≈0.58. The baseline ImageNet trained transformer model and TransMIL aggregation out-performed EfficientNet (macro-AUC=0.760, 95%CI 0.706-0.814) yet still trailed the stronger CNNs, particularly in class-balanced metrics (macro-F1 0.465 (95%CI 0.373-0.557), balanced accuracy 0.497 (95%CI 0.426-0.569)).

**Figure 2:**
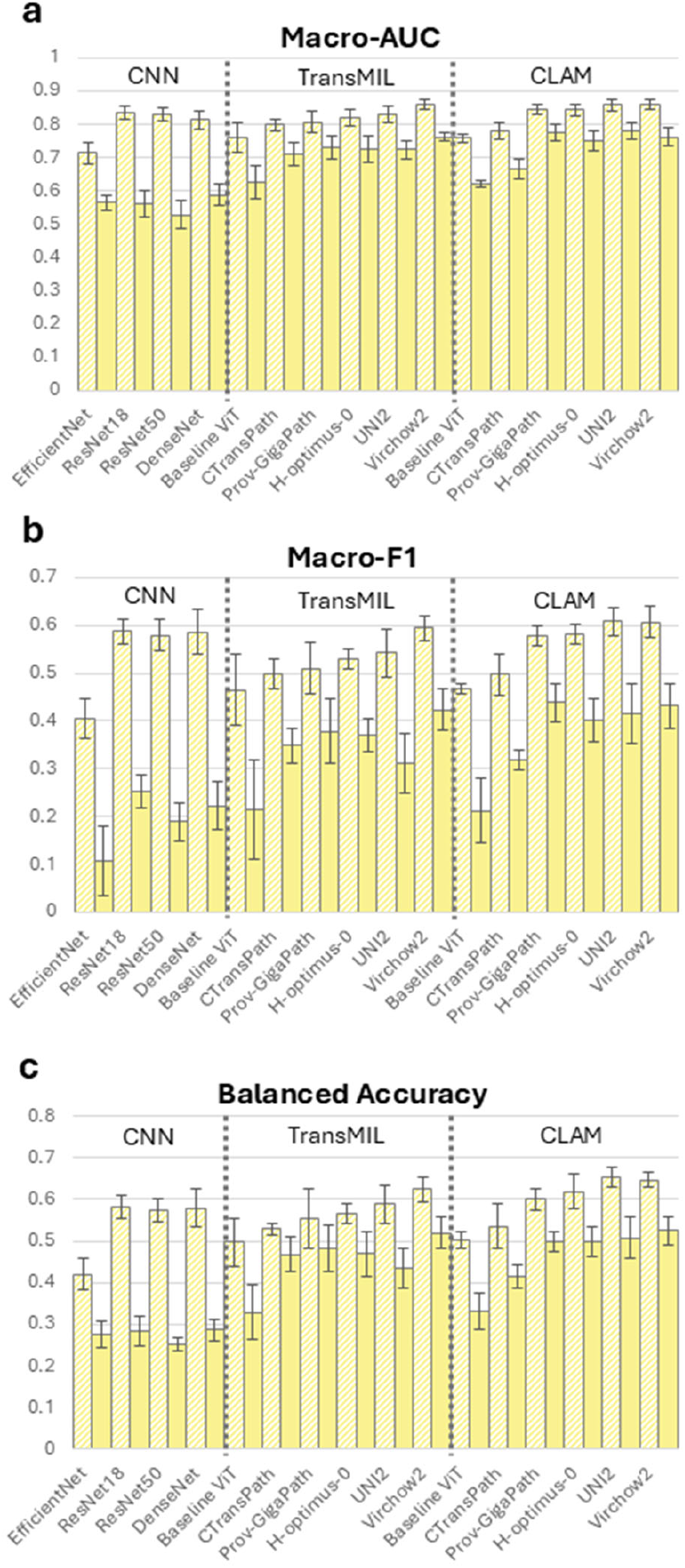
Model Macro-Metrics in Cross-Validation and External Validation. Comparison of (a) macro–area under the receiver operating characteristic curve (Macro-AUC), (b) macro–F1 score, and (c) balanced accuracy for CNN, TransMIL, and CLAM pipelines across foundation model feature extractors. Light bars represent mean performance across cross-validation folds and dark bars represent mean performance on external validation; error bars show standard deviation. Abbreviations: AUC, area under the curve; F1, F1 score; CLAM, clustering-constrained attention multiple instance learning; CNN, convolutional neural network; MIL, multiple instance learning.

Replacing the baseline ViT feature extractor with foundation encoders yielded a consistent improvement in performance. CTransPath, Prov-GigaPath, H-Optimus-0, UNI2 and Virchow2 with TransMIL aggregation all achieved higher macro-AUCs. Values ranged from 0.799 (95%CI 0.778-0.820) for CTransPath to 0.858 (95%CI 0.839-0.876) for Virchow2. Balanced accuracy climbed to 0.624 for Virchow2.

When the aggregation strategy was switched to the CLAM architecture, further improvements were observed for every backbone except baseline ViT. Prov-GigaPath, H-Optimus-0 and UNI2 saw the biggest improvements with CLAM over TransMIL (macro-AUC TransMIL to CLAM: 0.807 to 0.845, 0.820 to 0.844 and 0.830 to 0.858) with similar improvements in macro-F1 and balanced accuracy. Despite a lower absolute gain with the CLAM vs TransMIL architecture, the highest internal performance on cross-validation was Virchow2 with CLAM, which achieved a macro-AUC of 0.860 (95%CI 0.839-0.880), macro-F1 of 0.607 (95%CI 0.565-0.648) and a balanced accuracy of 0.647 (95%CI 0.623-0.670).

### External Validation

External validation on the independent cohort revealed a marked divergence between model architectures, Figure 2 (dark bars), Table S3. All CNNs experienced pronounced degradation in performance when compared to cross-validation: EfficientNet, DenseNet and both ResNets fell to macro-AUC of 0.528-0.588, macro-F1 below 0.252 and balanced accuracy less than 0.286.

The baseline ViT performed marginally better than CNNs with macro-AUC 0.626, this improvement was statistically significant in all comparisons except to DenseNet (p=0.130, Table S5). By contrast, foundation-model transformers retained substantially higher discrimination with TransMIL implementations producing macro-AUCs from 0.712-0.761, macro-F1s from 0.311-0.424 and balanced accuracies from 0.433-0.519. These were statistically superior to all CNNs and to the baseline ViT (Table S5).

The CLAM tile aggregation architecture again conferred an incremental benefit for every backbone with the exception of ViT and CTransPath, this difference was statistically significant in Prov-GigaPath and UNI2 (Table S5). In the external set, Prov-GigaPath with CLAM outperformed Prov-GigaPath with TransMIL by 0.046 macro-AUC points (0.777 vs 0.731) and achieved the highest macro-F1 of any model (0.438). UNI2 with CLAM yielded the top macro-AUC overall (0.780), with Virchow2 with CLAM close behind in macro-AUC (0.762) and best overall balanced accuracy (0.525).

### Subtype Performance

Across all four molecular subtypes, conventional CNN backbones performed reasonably well in cross-validation with AUCs reaching over 90% for p53abn and slightly lower for other subtypes, Figure 3, Table S2. However, in external validation, the CNNs had a dramatic fall off across all subtypes with AUCs seldom surpassing 0.60, Table S4. p53abn predictions were the highest with dMMR and POLE having very poor discrimination (~50%). The baseline ViT models had a similar trend with slightly better performance in external validation.

**Figure 3:**
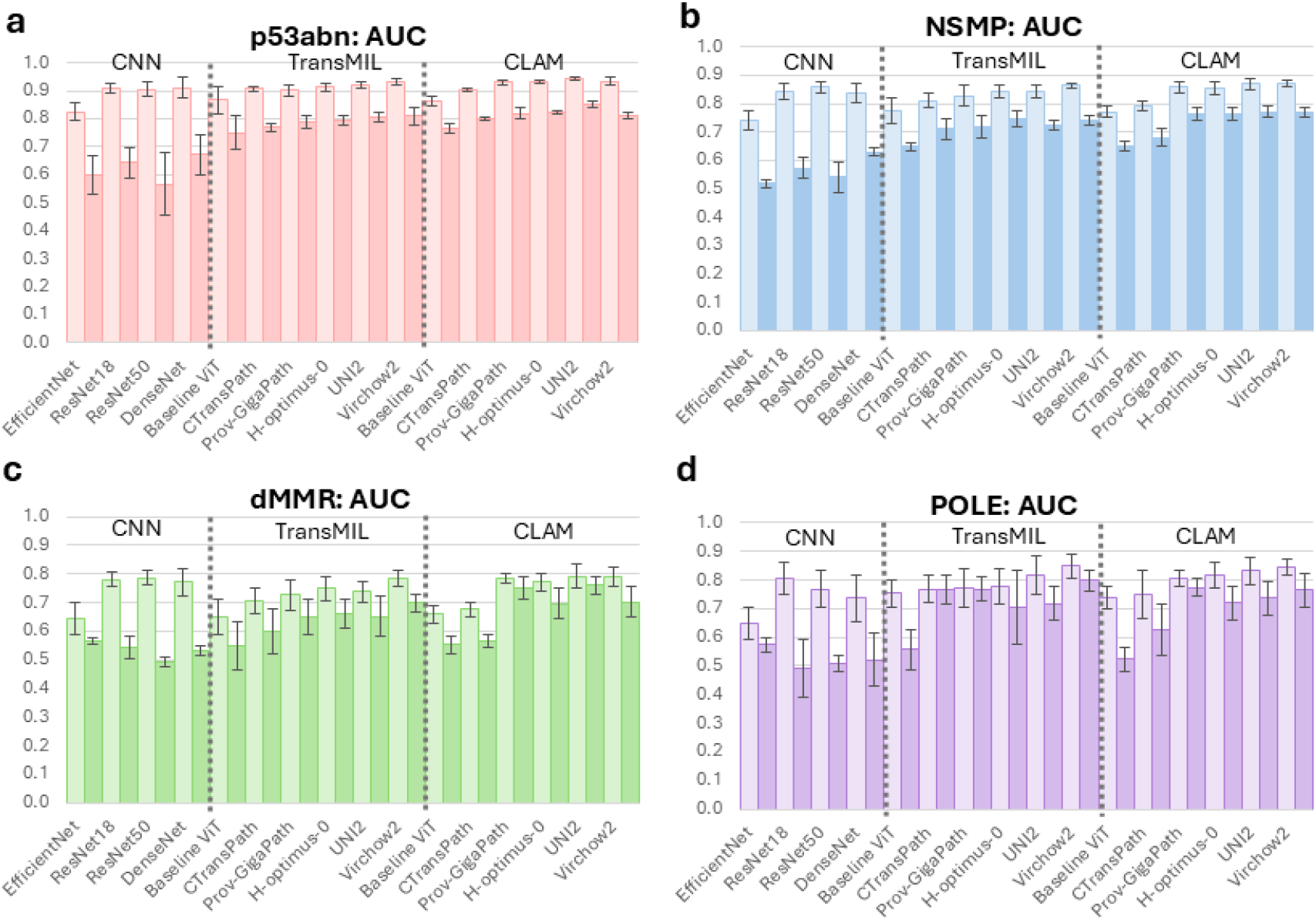
Model Subtype-AUC in Cross-Validation and External Validation. Subtype-level area under the receiver operating characteristic curve (AUCs) for (a) p53abn, (b) NSMP, (c) dMMR, and (d) POLE classifiers across CNN, TransMIL, and CLAM pipelines with different feature extractors. Light bars represent mean AUC across cross-validation folds and dark bars represent mean AUC from external validation; error bars show standard deviation. Abbreviations: AUC, area under the curve.

In contrast, models using foundation encoders showed consistently superior discrimination, Figure 3 (light bars), Tables S2. The choice of tile-aggregation strategy further had an effect performance. During cross-validation, UNI2 with CLAM had the highest performance for both p53abn and dMMR subtypes (AUC 0.942 (95%CI 0.933-0.951) and 0.792 (95%CI 0.757-0.827), respectively). Virchow2 with CLAM had the highest performance for NSMP (AUC 0.869 (95%CI 0.847-0.891)) but with TransMIL for POLE (AUC 0.849 (95%CI 0.810-0.888))

In external validation, UNI2 with CLAM had the highest AUC for all subtypes except POLE, Figure 3 (dark bars), Table S4. p53abn had the highest performance with an AUC of 0.851 (95%CI 0.837-0.865). NSMP and dMMR had AUC 0.770 (95%CI 0.746-0.794) and 0.759 (95%CI 0.718-0.800), respectively. Virchow2 with TransMIL had the highest prediction performance by AUC for POLE at 0.798 (95%CI 0.752-0.844). In most other cases, CLAM improved performance across subtypes when using foundation models.

### Interpretability Analysis - Qualitative

Review of attention maps revealed all models focused on tumor containing regions of the WSIs and largely ignored non-tumor regions, Figure 4. Expert pathologist review of high-scoring tiles revealed distinct morphologic correlates for each subtype.

**Figure 4:**
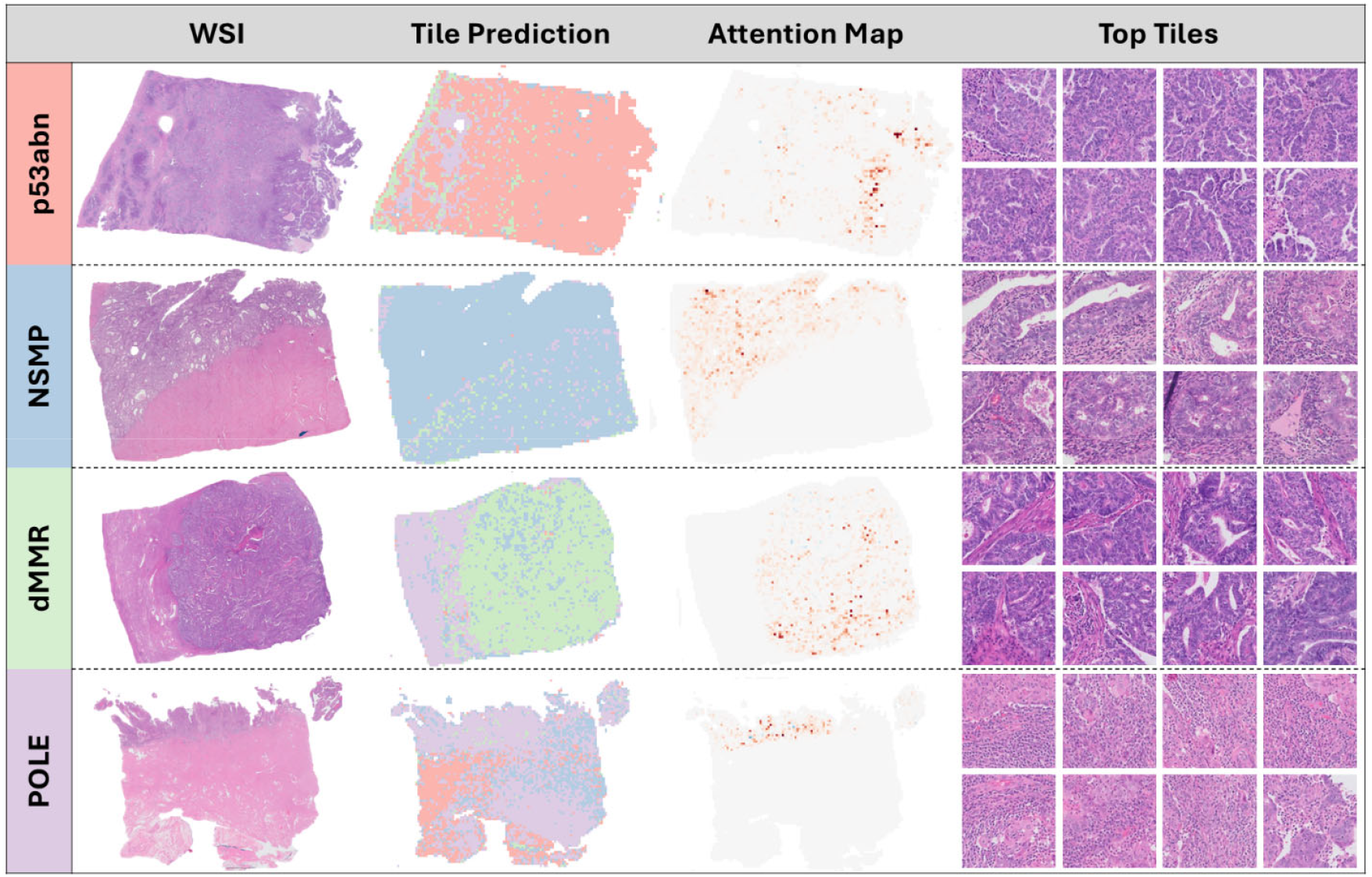
Whole Slide Image (WSI), Tiles and Attention Maps. Representative WSIs from each molecular subtype (rows) showing: (left to right) original WSI, tile-level subtype predictions, attention heatmaps, and the top tiles most predictive for that final predicted subtype. Tile prediction maps are color-coded by predicted class as seen along the left side; attention maps highlight regions contributing most to the slide-level prediction with darker areas receiving more attention. Abbreviations: WSI, whole-slide image.

p53abn: High nuclear-to-cytoplasmic (N/C) ratio with pleomorphic, hyperchromatic nuclei, coarse chromatin, prominent nucleoli, and frequent mitoses. Growth is papillary to micropapillary with irregular luminal contours and minimal inflammation.

NSMP: Low N/C ratio with bland, round nuclei and tubular glands, often with morular or tubal metaplasia. Stroma separates glands, luminal borders are smooth, and inflammation is variable but limited.

dMMR: Fused or cribriform glands with lumens, moderate N/C ratio, smooth luminal borders, and mild intra- and peritumoral lymphocytic inflammation without stromal desmoplasia.

POLE: Abundant peri- and intratumoral lymphoplasmacytic inflammation fragmenting tumor nests. Tumor shows low N/C ratio, smooth luminal borders when present, basal to pseudostratified nuclei, and prominent nucleoli but minimal atypia.

Overall, the models emphasized morphologic features consistent with expected subtype phenotypes: papillary/pleomorphic for p53abn, bland glandular for NSMP, intermediate glandular with mild inflammation for dMMR, and inflammation-dominant for POLE.

### Quantitative Analysis: Cell type composition

Based on quantitative evaluation using HoVerNet (Figure 5, Table S6), p53abn high prediction tiles are overwhelmingly tumor-cell–dominant: neoplastic 82% (highly statistically significant compared to all other subtypes, Table S7), with very little inflammatory (<2%), low stroma (<10%) and minimal necrosis (<2%). NSMP and POLE are markedly more stromal, each with >30% connective tissue and a lower tumor fraction (NSMP 60%, POLE 38%). dMMR sits between these extremes (neoplastic 73%, inflammatory 4.4%, connective 18%, dead <2%). dMMR tiles have a longer tail towards more inflammatory cells. POLE shows the most immune cells (16% inflammatory, highly statistically significant, Table S7) and the greatest necrosis (7.4% dead).

**Figure 5:**
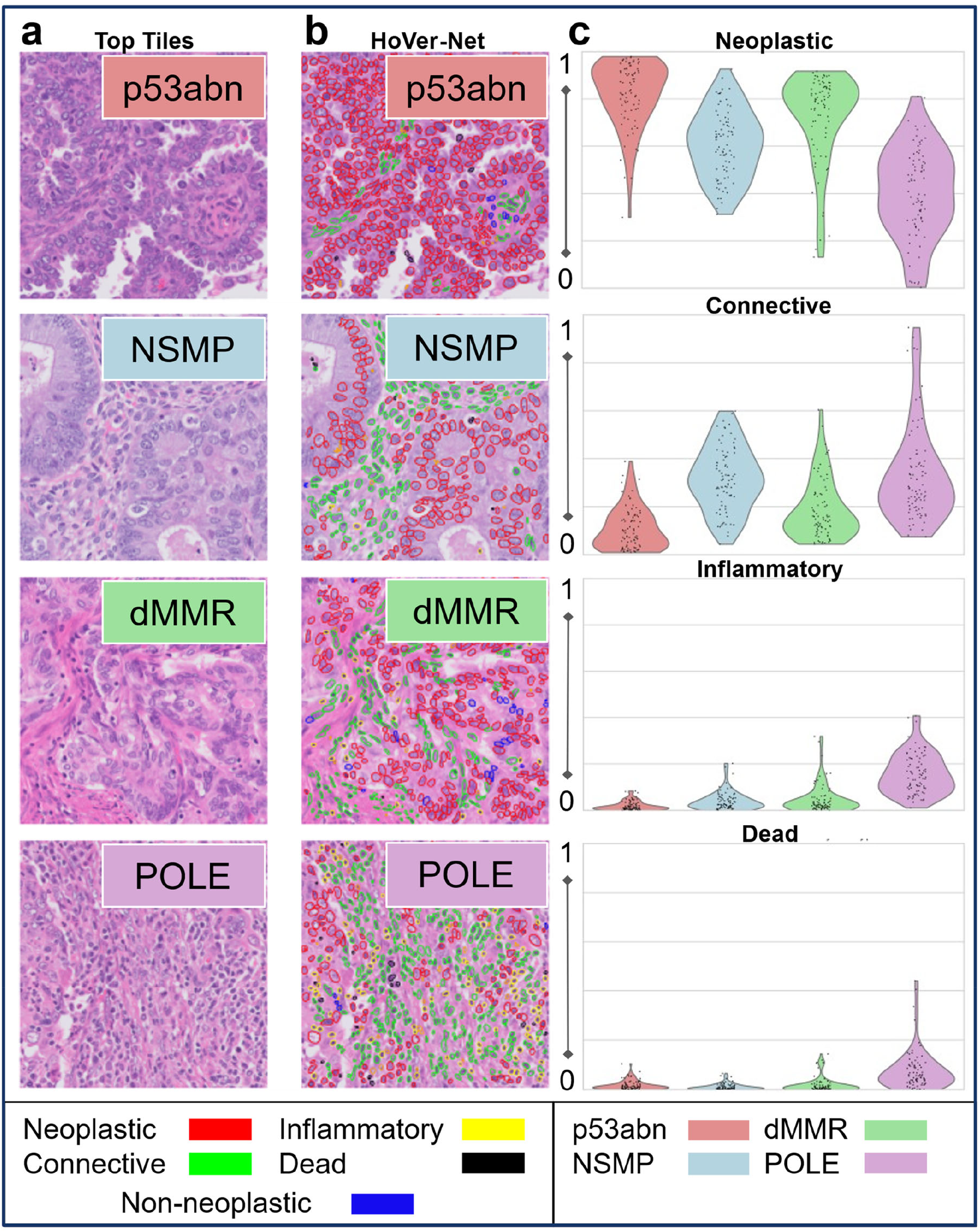
Interpretability Analysis by HoVer-Net. (a) Top tiles most predictive for each molecular subtype, (b) corresponding HoVer-Net nuclear segmentation maps, color-coded by cell type: neoplastic (red), connective (green), inflammatory (yellow), dead (black) and (c) violin plots plus jittered dot plot of per-tile proportion (0 to 1) showing the distribution of cell type fractions in top tiles across subtypes.

### Quantitative Analysis: Nuclear Morphometrics

Significant differences were identified in all nuclear morphometric metrics across the four molecular subtypes (Table S9, Figures S1A-S1E). Post-hoc Dunn’s tests with Holm correction revealed that mean nuclear area showed the most pronounced separation, with p53abn nuclei significantly larger than all other subtypes (Table S10). Circularity irregularity also demonstrated broad separation, with p53abn differing significantly from NSMP, dMMR, and POLE. Size dispersion and pleomorphism index primarily distinguished POLE from other subtypes, while eccentricity variability were limited to p53abn comparisons with NSMP and POLE. These findings suggest that both nuclear size and shape irregularity contribute to the morphological distinctiveness of certain subtypes, particularly p53abn.

## Discussion

In this study of 1,535 patients and 2,552 diagnostic H&E WSIs, we demonstrate that transformer-based pipelines built on open histopathology foundation encoders and trained on open-sourced data with attention-based MIL achieve state-of-the-art discrimination of molecular subtypes of endometrial cancer. The best configuration (UNI2 with CLAM) reached a macro-AUC of 0.86 in five-fold cross-validation and, critically, retained 0.78 macro-AUC on an independent patient cohort of real-world patients. Conventional ImageNet-pre-trained CNNs and an ImageNet-pre-trained ViT model suffered marked performance collapse on the same independent set, underscoring the importance of domain-specific pre-training and class-specific attention mechanisms for generalization. Foundation encoders capture histologic patterns that ImageNet-pretrained models miss. The foundation encoders allow more data efficiency when training so the MIL head can converge with hundreds rather than tens of thousands (or millions) of labeled slides. Furthermore, although all pipelines shared identical frozen foundation encoders, swapping TransMIL for the CLAM aggregator raised macro-AUC by 0.03-0.05 and boosted balanced accuracy by up to six percentage points on the independent cohort. We attribute this to CLAM-MB’s class-specific, instance-level attention, which explicitly compels the network to identify a small subset of diagnostically relevant tiles, an advantage with four subtypes and when endometrial-cancer slides contain extensive benign tissue. In contrast, TransMIL’s global self-attention, though expressive, appeared more prone to over-fitting. These findings highlight the importance of the aggregation architecture when translating AI models from training to real-world deployment.

p53abn consistently had the highest AUCs across configurations, this is most likely due to its overtly high-grade, tumor-dense morphology as noted in both qualitative and quantitative interpretation. NSMP also had consistent high performance across configurations and as it is the most common subtype, consistently had the highest overall accuracy. Both dMMR and POLE had high variation across configurations, although both performed reasonably well in our final model. This reflects their biologic heterogeneity and overlapping features with other subtypes. dMMR benefited disproportionately from CLAM. Misclassification patterns also track the histology (normalized confusion matrix, Figure S2): dMMR confused between NSMP and POLE likely owing to its intermediate architecture, cellular make-up and nuclear morphometrics, whereas POLE was most often incorrectly classified as p53abn or NSMP, consistent with its combination of pronounced heterogeneity on tile classification and wide variation of cellular content and nuclear morphometrics.

Several groups have previously published AI models for molecular classification in EC. Panoptes, a multi-resolution CNN based model, predicted driver mutations but lacked full subtyping validation^17^. im4MEC, developed by the PORTEC consortium, is a custom transformer-based model to classify subtypes in the PORTEC cohorts^18^. Other models, such as TR-MAMIL and MMRNet, focus on specific molecular features such as MMR status, MSI or tumor mutational burden^19, 20, 22^. hi-UNI, developed by Cui et al, introduced a hierarchical ViT backbone that leverages the foundation encoder UNI for subtype prediction^21^.

A distinctive advantage of our work is that every component of model development, from training WSIs and matched molecular labels to training pipeline and pre-trained encoders, is openly accessible. The discovery cohort was assembled exclusively from TCGA and CPTAC projects. All pre-processing and training code is released under an MIT license, and the foundation encoders are available for use (some with permission from their developers). In contrast, most other large studies that have developed prediction models were trained predominantly on private cohorts and therefore cannot be independently replicated or stress-tested^18, 21^. There was a previously reported model based on TCGA and CPTAC, however the model was built on a custom CNN without full validation on an external dataset so it is difficult to directly compare the performance^17^.

Open data and pipelines provide concrete benefits for this type of prediction model development. First, they enable external investigators to benchmark new models, encoders and aggregation schemes or fine-tune for rarer entities without starting from scratch. Second, regulators now expect traceability for AI-based diagnostics; open source shortens the path to FDA clearance and adoption. Finally, transparent resources foster multicenter collaboration, essential for ensuring generalizability which is especially important for more rare subtypes. Because we used the publicly available data for training, we were able to add a more stringent external validation, rather than validating on TCGA, the performance of our model was measured on a real-world 720 patient cohort providing a realistic gauge of deployment readiness.

Additional strengths include our interpretability analysis, which combines qualitative review (providing face validity for pathologists) with quantitative metrics (offering reproducible, model-agnostic evidence aligned with known biology). This dual approach supports both clinical trust and benchmarking. We also conducted three-tier evaluation (development, cross-validation, external validation) consistent with TRIPOD guidelines, using rigorous statistics to minimize bias^43^. Limitations include retrospective slide collections; scanner and staining variability (partially mitigated by the external cohort); and class imbalance. Subtype distributions also differed across cohorts. Attention maps are not causal explanations but pairing them with quantitative analyses reduces over-interpretation and strengthens validity. Finally, our validated cohort may not fully reflect all deployment settings.

With the increasing adoption of molecular classification in the clinical management of EC, limitations of traditional histopathology, including interobserver variability, as well as the cost, time, and resources required for comprehensive molecular testing highlight the need for scalable, high-throughput alternatives. Our findings demonstrate that transformer-based AI models applied to routine H&E slides can accurately predict molecular subtypes of endometrial cancer, providing evidence that an AI-driven platform can serve as a surrogate for molecular classification. Such an approach has the potential to function as a clinical decision-support tool, either by prompting reflex confirmatory testing or by serving as a practical alternative in low-resource settings.

Ultimately, this strategy could broaden access to molecularly informed care and enable more equitable implementation of precision oncology for EC worldwide.

## Data, Code and Model Availability

### Data availability

Digital histopathology slides from our institutional clinical cohort contain protected health information and are covered by HIPAA and our IRB; these data cannot be posted publicly. De-identified derivatives (e.g. tile-level features, slide-level predictions, and heatmaps) may be shared under a data use agreement upon reasonable request to the corresponding author and pending institutional approvals. Public datasets used in this study are available from TCGA via the NCI Genomic Data Commons (https://portal.gdc.cancer.gov) and from CPTAC via The Cancer Imaging Archive (https://www.cancerimagingarchive.net).

### Code availability

Analyses were performed using open-source frameworks; repositories are publicly available at: STAMP (https://github.com/KatherLab/STAMP), HIA (https://github.com/KatherLab/HIA), and CLAM (https://github.com/mahmoodlab/CLAM). Any custom scripts used to orchestrate experiments are available from the corresponding author on reasonable request.

### Model availability

Foundation models referenced in this work are openly available at:

- ViT-Base (https://huggingface.co/google/vit-base-patch16-224-in21k)
- CTransPath (https://huggingface.co/jamesdolezal/CTransPath)
- Prov-GigaPath (https://huggingface.co/prov-gigapath/prov-gigapath)
- H-Optimus-0 (https://huggingface.co/bioptimus/H-optimus-0)
- UNI-2-h (https://huggingface.co/MahmoodLab/UNI2-h)
- Virchow2 (https://huggingface.co/paige-ai/Virchow2)

### Software tools

Additional tools used include:

- TIA Toolbox v1.0.0 (https://tiatoolbox.readthedocs.io/en/v1.0.0/index.html)
- HoVer-Net (https://github.com/vqdang/hover_net)

## Author Contributions

VMW led conceptualization, methodology, and software development, performed formal analyses, and drafted the original manuscript. CMC contributed to conceptualization and resources and assisted with review and editing. SJC and MIS performed pathology review; DTG performed pathology review and contributed to review and editing. MJG provided supervision and resources. JGB contributed to conceptualization, supervision, and manuscript review and editing. All authors reviewed and approved the final manuscript.

## Acknowledgements

None

## Disclaimers

None, the authors declare no potential conflicts of interest.

