## Supplementary Appendix 1 for "Real-World Benchmarking and Validation of Foundation Model Transformers for Endometrial Cancer Subtyping from Histopathology"

**Supplemental Appendix 1**

Table S1: Cross-validation metrics including Macro-area under the receiver operating characteristic curve (AUC), macro-F1 score and balanced accuracy, all with 95% confidence interval for each feature extractor and aggregation type.

Table S2: Cross-validation area under the receiver operating characteristic curve (AUC) by subtype with 95% confidence interval for each feature extractor and aggregation type.

Table S3: External validation metrics including Macro-area under the receiver operating characteristic curve (AUC), macro-F1 score and balanced accuracy, all with 95% confidence interval for each feature extractor and aggregation type.

Table S4: External validation area under the receiver operating characteristic curve (AUC) by subtype with 95% confidence interval for each feature extractor and aggregation type.

Table S5: Pairwise model comparisons of AUC performance: Differences in area under the receiver operating characteristic curve (ΔAUC) between model pairs are shown with corresponding 95% confidence intervals (CI Low, CI High), raw p values, Shapiro–Wilk normality test results, and Holm-adjusted p values for multiple comparisons. Positive ΔAUC indicates superior performance of Model 1 over Model 2. Abbreviations: AUC, area under the curve; CI, confidence interval; CNN, convolutional neural network; ViT, vision transformer; CLAM, clustering-constrained attention multiple instance learning; TransMIL, transformer-based multiple instance learning.

Table S6: HoVerNet nuclear segmentation results by molecular subtype, all given as percentage of total cells per tile, averaged across top tiles.

Table S7: Kruskal–Wallis test results for cell-type composition across molecular subtypes: Non-parametric Kruskal–Wallis tests were used to assess differences in cell-type fractions between molecular subtypes. For each cell type, the H statistic, p value, effect size (ε²), and total number of observations (N) are reported. Abbreviations: ε², epsilon-squared effect size; N, number of observations. Scientific notation (E) indicates ×10^power.

Table S8: Dunn pairwise comparisons of cell-type composition between molecular subtypes with Holm correction. Dunn’s post hoc tests were performed following significant Kruskal–Wallis results to compare cell-type fractions between all pairs of molecular subtypes. Only significant results are listed. Raw p values and Holm-adjusted p values for multiple comparisons are reported. Scientific notation (E) indicates ×10^power.

Table S9: Kruskal–Wallis test results for nuclear morphometric testing across molecular subtypes: Non-parametric Kruskal–Wallis tests were used to assess differences in nuclear morphometrics between molecular subtypes. For each cell type, the H statistic, p value, effect size (ε²), and total number of observations (N) are reported. Abbreviations: ε², epsilon-squared effect size; N, number of observations. Scientific notation (E) indicates ×10^power.

Table S10: Dunn pairwise comparisons of nuclear morphometrics inculding mean nuclear area, coefficient of variation of area, pleomorphism index, coefficient of variation of eccentricity and coefficient of variation of circularity between molecular subtypes with Holm correction. Dunn’s post hoc tests were performed following significant Kruskal–Wallis results to compare between all pairs of molecular subtypes. Only significant results are listed. Raw p values and Holm-adjusted p values for multiple comparisons are reported. Scientific notation (E) indicates ×10^power.

Figure S1: Violin plots of nuclear morphometrics across subtypes, (A) Mean area, (B) coefficient of variation of area, (C) pleomorphism index, (D) coefficient of variation of eccentricity and (E) coefficient of variation of circularity. Subtypes are CNV-H (p53abn), CNV-L (NSMP), MSI-H (dMMR) and POLE.

Figure S2: Normalized confusion matrix for UNI2 with CLAM: Confusion matrix showing classification performance of the UNI2 vision transformer with CLAM aggregation for predicting molecular subtypes. Values represent row-normalized proportions of true subtype cases (rows) predicted as each subtype (columns). Darker colors indicate higher proportions.

Table S1: Cross-validation metrics including Macro-area under the receiver operating characteristic curve (AUC), macro-F1 score and balanced accuracy, all with 95% confidence interval for each feature extractor and aggregation type.

| **5-Fold Cross-Validation** | | | |
| --- | --- | --- | --- |
| **Model** | **Macro-AUC (95%CI)** | **Macro-F1 (95%CI)** | **Balanced accuracy (95%CI)** |
| EfficientNet | 0.715 (0.675-0.754) | 0.404 (0.353-0.455) | 0.420 (0.372-0.468) |
| ResNet-18 | 0.834 (0.810-0.859) | 0.587 (0.555-0.619) | 0.582 (0.549-0.616) |
| ResNet-50 | 0.829 (0.803-0.854) | 0.580 (0.537-0.622) | 0.574 (0.540-0.608) |
| DenseNet | 0.813 (0.781-0.845) | 0.585 (0.528-0.643) | 0.580 (0.524-0.635) |
| Baseline ViT (TransMIL) | 0.760 (0.706-0.814) | 0.465 (0.373-0.557) | 0.497 (0.426-0.569) |
| Baseilne ViT (CLAM) | 0.758 (0.740-0.776) | 0.467 (0.446-0.490) | 0.504 (0.480-0.528) |
| CTransPath (TransMIL) | 0.799 (0.778-0.820) | 0.498 (0.468-0.528) | 0.529 (0.514-0.544) |
| CTransPath (CLAM) | 0.780 (0.748-0.812) | 0.498 (0.454-0.542) | 0.536 (0.490-0.582) |
| Prov-GigaPath (TransMIL) | 0.807 (0.773-0.842) | 0.510 (0.448-0.572) | 0.555 (0.504-0.606) |
| Prov-GigaPath (CLAM) | 0.845 (0.822-0.867) | 0.578 (0.556-0.600) | 0.600 (0.575-0.625) |
| H-Optimus-0 (TransMIL) | 0.820 (0.792-0.848) | 0.528 (0.509-0.548) | 0.567 (0.541-0.592) |
| H-Optimus-0 (CLAM) | 0.844 (0.822-0.866) | 0.582 (0.556-0.609) | 0.620 (0.567-0.672) |
| UNI2 (TransMIL) | 0.830 (0.796-0.864) | 0.542 (0.481-0.604) | 0.589 (0.532-0.647) |
| UNI2 (CLAM) | 0.858 (0.836-0.881) | 0.608 (0.570-0.646) | 0.654 (0.626-0.681) |
| Virchow2 (TransMIL) | 0.858 (0.839-0.876) | 0.594 (0.562-0.627) | 0.624 (0.586-0.662) |
| Virchow2 (CLAM) | 0.860 (0.839-0.880) | 0.607 (0.565-0.648) | 0.647 (0.623-0.670) |

Table S2: Cross-validation area under the receiver operating characteristic curve (AUC) by subtype with 95% confidence interval for each feature extractor and aggregation type.

| **5-Fold Cross-Validation (Subtypes)** | | | | |
| --- | --- | --- | --- | --- |
| **Model** | **NSMP AUC (95%CI)** | **p53abn AUC (95%CI)** | **dMMR AUC (95%CI)** | **POLE AUC (95%CI)** |
| EfficientNet | 0.739 (0.698-0.780) | 0.823 (0.783-0.863) | 0.645 (0.577-0.713) | 0.651 (0.581-0.721) |
| ResNet-18 | 0.842 (0.810-0.874) | 0.908 (0.888-0.928) | 0.781 (0.746-0.816) | 0.806 (0.733-0.879) |
| ResNet-50 | 0.857 (0.833-0.881) | 0.904 (0.872-0.936) | 0.785 (0.753-0.817) | 0.769 (0.691-0.847) |
| DenseNet | 0.836 (0.793-0.879) | 0.909 (0.863-0.955) | 0.770 (0.714-0.826) | 0.736 (0.634-0.838) |
| Baseline ViT (TransMIL) | 0.773 (0.718-0.828) | 0.866 (0.806-0.926) | 0.648 (0.571-0.725) | 0.753 (0.691-0.815) |
| Baseline ViT (CLAM) | 0.772 (0.744-0.800) | 0.864 (0.847-0.881) | 0.659 (0.627-0.691) | 0.739 (0.699-0.779) |
| CTransPath (TransMIL) | 0.811 (0.778-0.844) | 0.907 (0.888-0.926) | 0.708 (0.648-0.768) | 0.769 (0.706-0.832) |
| CTransPath (CLAM) | 0.792 (0.764-0.820) | 0.904 (0.888-0.920) | 0.676 (0.642-0.710) | 0.750 (0.612-0.888) |
| Prov-GigaPath (TransMIL) | 0.828 (0.784-0.872) | 0.900 (0.882-0.918) | 0.726 (0.663-0.789) | 0.772 (0.700-0.844) |
| Prov-GigaPath (CLAM) | 0.859 (0.836-0.882) | 0.930 (0.919-0.941) | 0.785 (0.764-0.806) | 0.807 (0.779-0.835) |
| H-Optimus-0 (TransMIL) | 0.844 (0.812-0.876) | 0.912 (0.893-0.931) | 0.748 (0.688-0.808) | 0.777 (0.710-0.844) |
| H-Optimus-0 (CLAM) | 0.853 (0.826-0.880) | 0.932 (0.924-0.940) | 0.771 (0.739-0.803) | 0.818 (0.779-0.857) |
| UNI2 (TransMIL) | 0.843 (0.811-0.875) | 0.921 (0.910-0.932) | 0.738 (0.693-0.783) | 0.817 (0.737-0.897) |
| UNI2 (CLAM) | 0.868 (0.845-0.891) | 0.942 (0.933-0.951) | 0.792 (0.757-0.827) | 0.831 (0.792-0.870) |
| Virchow2 (TransMIL) | 0.863 (0.848-0.878) | 0.933 (0.923-0.943) | 0.785 (0.753-0.817) | 0.849 (0.810-0.888) |
| Virchow2 (CLAM) | 0.869 (0.847-0.891) | 0.933 (0.910-0.956) | 0.790 (0.756-0.824) | 0.847 (0.818-0.876) |

Table S3: External validation metrics including Macro-area under the receiver operating characteristic curve (AUC), macro-F1 score and balanced accuracy, all with 95% confidence interval for each feature extractor and aggregation type.

| **External Validation** | | | |
| --- | --- | --- | --- |
| **Model** | **Macro-AUC (95%CI)** | **Macro-F1 (95%CI)** | **Balanced accuracy (95%CI)** |
| EfficientNet | 0.564 (0.535-0.593) | 0.106 (0.015-0.196) | 0.274 (0.235-0.313) |
| ResNet-18 | 0.563 (0.512-0.613) | 0.252 (0.208-0.296) | 0.283 (0.238-0.327) |
| ResNet-50 | 0.528 (0.475-0.581) | 0.189 (0.140-0.238) | 0.253 (0.233-0.273) |
| DenseNet | 0.588 (0.546-0.630) | 0.223 (0.160-0.286) | 0.286 (0.255-0.317) |
| ViT (TransMIL) | 0.626 (0.576-0.675) | 0.215 (0.063-0.367) | 0.329 (0.277-0.381) |
| ViT (CLAM) | 0.622 (0.607-0.637) | 0.212 (0.127-0.298) | 0.331 (0.299-0.364) |
| CTransPath (TransMIL) | 0.712 (0.680-0.745) | 0.348 (0.318-0.379) | 0.468 (0.437-0.499) |
| CTransPath (CLAM) | 0.667 (0.636-0.698) | 0.319 (0.302-0.336) | 0.416 (0.388-0.444) |
| Prov-GigaPath (TransMIL) | 0.731 (0.698-0.764) | 0.378 (0.297-0.459) | 0.482 (0.438-0.526) |
| Prov-GigaPath (CLAM) | 0.777 (0.753-0.801) | 0.438 (0.402-0.474) | 0.500 (0.475-0.525) |
| H-Optimus-0 (TransMIL) | 0.727 (0.680-0.774) | 0.369 (0.330-0.408) | 0.469 (0.425-0.513) |
| H-Optimus-0 (CLAM) | 0.750 (0.721-0.779) | 0.401 (0.361-0.441) | 0.498 (0.468-0.529) |
| UNI2 (TransMIL) | 0.724 (0.691-0.757) | 0.311 (0.233-0.389) | 0.433 (0.374-0.493) |
| UNI2 (CLAM) | 0.780 (0.750-0.810) | 0.416 (0.339-0.493) | 0.507 (0.445-0.570) |
| Virchow2 (TransMIL) | 0.761 (0.745-0.778) | 0.424 (0.370-0.478) | 0.519 (0.472-0.566) |
| Virchow2 (CLAM) | 0.762 (0.730-0.794) | 0.431 (0.371-0.490) | 0.525 (0.484-0.567) |

Table S4: External validation area under the receiver operating characteristic curve (AUC) by subtype with 95% confidence interval for each feature extractor and aggregation type.

| **External Validation (Subtypes)** | | | | |
| --- | --- | --- | --- | --- |
| **Model** | **NSMP AUC (95%CI)** | **p53abn AUC (95%CI)** | **dMMR AUC (95%CI)** | **POLE AUC (95%CI)** |
| EfficientNet | 0.518 (0.498-0.538) | 0.600 (0.516-0.684) | 0.563 (0.548-0.578) | 0.575 (0.544-0.606) |
| ResNet-18 | 0.573 (0.528-0.618) | 0.643 (0.575-0.711) | 0.541 (0.491-0.591) | 0.493 (0.369-0.617) |
| ResNet-50 | 0.540 (0.470-0.610) | 0.567 (0.429-0.705) | 0.493 (0.471-0.515) | 0.510 (0.474-0.546) |
| DenseNet | 0.630 (0.614-0.646) | 0.671 (0.584-0.758) | 0.531 (0.509-0.553) | 0.520 (0.405-0.635) |
| ViT (TransMIL) | 0.648 (0.633-0.663) | 0.750 (0.676-0.824) | 0.548 (0.446-0.650) | 0.557 (0.471-0.643) |
| ViT (CLAM) | 0.650 (0.630-0.670) | 0.763 (0.741-0.785) | 0.554 (0.516-0.592) | 0.523 (0.472-0.574) |
| CTransPath (TransMIL) | 0.711 (0.666-0.756) | 0.768 (0.749-0.787) | 0.600 (0.504-0.696) | 0.768 (0.703-0.833) |
| CTransPath (CLAM) | 0.681 (0.640-0.722) | 0.798 (0.791-0.805) | 0.565 (0.539-0.591) | 0.625 (0.515-0.735) |
| Prov-GigaPath (TransMIL) | 0.717 (0.669-0.765) | 0.789 (0.760-0.818) | 0.649 (0.573-0.725) | 0.769 (0.714-0.824) |
| Prov-GigaPath (CLAM) | 0.765 (0.738-0.792) | 0.818 (0.793-0.843) | 0.749 (0.699-0.799) | 0.775 (0.735-0.815) |
| H-Optimus-0 (TransMIL) | 0.746 (0.712-0.780) | 0.794 (0.775-0.813) | 0.662 (0.599-0.725) | 0.706 (0.546-0.866) |
| H-Optimus-0 (CLAM) | 0.762 (0.733-0.791) | 0.822 (0.817-0.827) | 0.697 (0.630-0.764) | 0.720 (0.648-0.792) |
| UNI2 (TransMIL) | 0.723 (0.703-0.743) | 0.804 (0.782-0.826) | 0.651 (0.565-0.737) | 0.717 (0.645-0.789) |
| UNI2 (CLAM) | 0.770 (0.746-0.794) | 0.851 (0.837-0.865) | 0.759 (0.718-0.800) | 0.738 (0.665-0.811) |
| Virchow2 (TransMIL) | 0.740 (0.718-0.762) | 0.809 (0.772-0.846) | 0.699 (0.659-0.739) | 0.798 (0.752-0.844) |
| Virchow2 (CLAM) | 0.768 (0.747-0.789) | 0.812 (0.791-0.833) | 0.703 (0.636-0.770) | 0.765 (0.691-0.839) |

Table S5: Pairwise model comparisons of AUC performance: Differences in area under the receiver operating characteristic curve (ΔAUC) between model pairs are shown with corresponding 95% confidence intervals (CI Low, CI High), raw p values, Shapiro–Wilk normality test results, and Holm-adjusted p values for multiple comparisons. Positive ΔAUC indicates superior performance of Model 1 over Model 2. Abbreviations: AUC, area under the curve; CI, confidence interval; CNN, convolutional neural network; ViT, vision transformer; CLAM, clustering-constrained attention multiple instance learning; TransMIL, transformer-based multiple instance learning.

| **Model 1** | **Model 2** | **ΔAUC** | **CI (Low)** | **CI (High)** | **p Value (Raw)** | **Shapiro-Wilk** | **p Value (Adjusted)** |
| --- | --- | --- | --- | --- | --- | --- | --- |
| EFFICIENTNET | baseline ViT (transMIL) | -0.058 | -0.085 | -0.031 | 0.004 | 0.921 | ***0.007*** |
| RESNET18 | baseline ViT (transMIL) | -0.060 | -0.106 | -0.013 | 0.024 | 0.971 | ***0.032*** |
| RESNET50 | baseline ViT (transMIL) | -0.095 | -0.142 | -0.048 | 0.005 | 0.598 | ***0.008*** |
| DENSENET | baseline ViT (transMIL) | -0.034 | -0.081 | 0.012 | 0.112 | 0.411 | 0.130 |
| CTransPath (TransMIL) | baseline ViT (transMIL) | 0.045 | 0.013 | 0.076 | 0.017 | 0.542 | ***0.024*** |
| H-Optimus-0 (TransMIL) | baseline ViT (transMIL) | 0.128 | 0.101 | 0.155 | 0.000 | 0.424 | ***0.001*** |
| Prov-GigaPath (TransMIL) | baseline ViT (transMIL) | 0.154 | 0.127 | 0.182 | 0.000 | 0.504 | ***0.001*** |
| UNI2 (TransMIL) | baseline ViT (transMIL) | 0.157 | 0.140 | 0.175 | 0.000 | 0.836 | ***0.000*** |
| Virchow2 (TransMIL) | baseline ViT (transMIL) | 0.140 | 0.113 | 0.167 | 0.000 | 0.276 | ***0.001*** |
| CTransPath (TransMIL) | DENSENET | 0.079 | 0.021 | 0.137 | 0.019 | 0.479 | ***0.027*** |
| CTransPath (TransMIL) | H-Optimus-0 (TransMIL) | -0.083 | -0.121 | -0.045 | 0.004 | 0.110 | ***0.007*** |
| CTransPath (TransMIL) | Prov-GigaPath (TransMIL) | -0.110 | -0.162 | -0.057 | 0.004 | 0.655 | ***0.007*** |
| CTransPath (TransMIL) | UNI2 (TransMIL) | -0.113 | -0.138 | -0.087 | 0.000 | 0.446 | ***0.001*** |
| CTransPath (TransMIL) | Virchow2 (TransMIL) | -0.095 | -0.129 | -0.060 | 0.002 | 0.580 | ***0.003*** |
| UNI2 (TransMIL) | Virchow2 (TransMIL) | 0.018 | -0.014 | 0.050 | 0.197 | 0.773 | 0.222 |
| Prov-GigaPath (TransMIL) | Virchow2 (TransMIL) | 0.015 | -0.028 | 0.057 | 0.394 | 0.156 | 0.410 |
| Prov-GigaPath (TransMIL) | UNI2 (TransMIL) | -0.003 | -0.043 | 0.037 | 0.840 | 0.427 | 0.856 |
| H-Optimus-0 (TransMIL) | Virchow2 (TransMIL) | -0.012 | -0.043 | 0.020 | 0.361 | 0.955 | 0.382 |
| H-Optimus-0 (TransMIL) | UNI2 (TransMIL) | -0.029 | -0.060 | 0.001 | 0.055 | 0.306 | 0.068 |
| H-Optimus-0 (TransMIL) | Prov-GigaPath (TransMIL) | -0.026 | -0.051 | -0.002 | 0.041 | 0.898 | 0.052 |
| Baseline ViT (CLAM) | baseline ViT (transMIL) | -0.003 | -0.063 | 0.057 | 0.887 | 0.687 | 0.900 |
| CTransPath (CLAM) | CTransPath (TransMIL) | -0.045 | -0.081 | -0.008 | 0.027 | 0.294 | 0.052 |
| H-Optimus-0 (CLAM) | H-Optimus-0 (TransMIL) | 0.023 | -0.032 | 0.078 | 0.302 | 0.647 | 0.350 |
| Prov-GigaPath (CLAM) | Prov-GigaPath (TransMIL) | 0.046 | 0.023 | 0.068 | 0.005 | 0.860 | ***0.013*** |
| UNI2 (CLAM) | UNI2 (TransMIL) | 0.056 | 0.028 | 0.084 | 0.005 | 0.881 | ***0.013*** |
| Virchow2 (CLAM) | Virchow2 (TransMIL) | 0.001 | -0.025 | 0.026 | 0.949 | 0.129 | 0.949 |

Table S6: HoVerNet nuclear segmentation results by molecular subtype, all given as percentage of total cells per tile, averaged across top tiles.

| **HoVerNet – Cell Types by Subtype** | | | | | |
| --- | --- | --- | --- | --- | --- |
|  | **Neoplastic epithelial** | **Inflammatory** | **Connective** | **Dead** | **Non-Neoplastic epithelial** |
| **p53abn** | 82.11% | 1.84% | 9.68% | 1.51% | 3.69% |
| **NSMP** | 59.90% | 4.21% | 30.73% | 1.13% | 2.27% |
| **dMMR** | 72.71% | 4.40% | 17.52% | 1.59% | 2.18% |
| **POLE** | 38.46% | 16.15% | 32.36% | 7.43% | 3.26% |

Table S7: Kruskal–Wallis test results for cell-type composition across molecular subtypes: Non-parametric Kruskal–Wallis tests were used to assess differences in cell-type fractions between molecular subtypes. For each cell type, the H statistic, p value, effect size (ε²), and total number of observations (N) are reported. Abbreviations: ε², epsilon-squared effect size; N, number of observations. Scientific notation (E) indicates ×10^power.

| **Kruskal–Wallis Testing – Cell Types** | | | | |
| --- | --- | --- | --- | --- |
| **Cell Type** | **H Statistic** | **p value** | **ε² (effect size)** | **N** |
| **Neoplastic epithelial** | 86.73199373 | 1.10E-18 | 0.264974664 | 320 |
| **Inflammatory** | 149.2838526 | 3.76E-32 | 0.462923584 | 320 |
| **Connective** | 144.5423506 | 3.96E-31 | 0.447918831 | 320 |
| **Dead** | 96.71583313 | 7.90E-21 | 0.296569092 | 320 |
| **Non-Neoplastic epithelial** | 17.00767501 | 7.04E-04 | 0.044328085 | 320 |

Table S8: Dunn pairwise comparisons of cell-type composition between molecular subtypes with Holm correction. Dunn’s post hoc tests were performed following significant Kruskal–Wallis results to compare cell-type fractions between all pairs of molecular subtypes. Only significant results are listed. Raw p values and Holm-adjusted p values for multiple comparisons are reported. Scientific notation (E) indicates ×10^power.

| **Dunn-Pairwise Test with Holm Correction – Cell Types** | | | | |
| --- | --- | --- | --- | --- |
| **Cell Type** | **Group 1** | **Group 2** | **p value (Raw)** | **p value (Holm)** |
| Neoplastic epithelial | P53ABN | POLE | 3.81E-15 | 2.28E-14 |
| Neoplastic epithelial | DMMR | POLE | 1.04E-14 | 5.18E-14 |
| Neoplastic epithelial | NSMP | POLE | 1.86E-08 | 7.46E-08 |
| Neoplastic epithelial | P53ABN | NSMP | 8.45E-04 | 2.53E-03 |
| Neoplastic epithelial | NSMP | DMMR | 3.04E-03 | 6.07E-03 |
| Inflammatory | P53ABN | POLE | 3.65E-25 | 2.19E-24 |
| Inflammatory | DMMR | POLE | 1.13E-17 | 5.64E-17 |
| Inflammatory | NSMP | POLE | 2.25E-17 | 9.02E-17 |
| Inflammatory | P53ABN | NSMP | 7.10E-07 | 1.45E-06 |
| Inflammatory | P53ABN | DMMR | 4.85E-07 | 1.45E-06 |
| Connective | P53ABN | NSMP | 2.44E-21 | 1.47E-20 |
| Connective | P53ABN | POLE | 8.26E-21 | 4.13E-20 |
| Connective | NSMP | DMMR | 1.46E-10 | 5.85E-10 |
| Connective | DMMR | POLE | 5.67E-10 | 1.70E-09 |
| Connective | P53ABN | DMMR | 3.29E-09 | 6.58E-09 |
| Dead | NSMP | POLE | 2.25E-16 | 1.35E-15 |
| Dead | P53ABN | POLE | 3.67E-15 | 1.84E-14 |
| Dead | DMMR | POLE | 1.95E-14 | 7.82E-14 |
| Non-Neoplastic epithelial | P53ABN | POLE | 3.46E-05 | 2.08E-04 |
| Non-Neoplastic epithelial | P53ABN | NSMP | 9.27E-03 | 4.64E-02 |

Table S9: Kruskal–Wallis test results for nuclear morphometric testing across molecular subtypes: Non-parametric Kruskal–Wallis tests were used to assess differences in nuclear morphometrics between molecular subtypes. For each cell type, the H statistic, p value, effect size (ε²), and total number of observations (N) are reported. Abbreviations: ε², epsilon-squared effect size; N, number of observations. Scientific notation (E) indicates ×10^power.

| **Kruskal–Wallis Testing – Nuclear Morphometrics** | | | | |
| --- | --- | --- | --- | --- |
| **Metric** | **H Statistic** | **p value** | **ε² (effect size)** | **N** |
| **Mean nuclear area** | 132.8411 | 3.1E-29 | 0.41089 | 320 |
| **Coefficient of variation of area** | 28.67438 | 3.00E-06 | 0.081248 | 320 |
| **Pleomorphism index (PI)** | 16.63818 | 8.39E-04 | 0.043159 | 320 |
| **Coefficient of variation of eccentricity** | 10.6877 | 0.01354 | 0.024328 | 320 |
| **Coefficient of variation of circularity** | 59.30116 | 3.4E-13 | 0.178168 | 320 |

Table S10: Dunn pairwise comparisons of nuclear morphometrics inculding mean nuclear area, coefficient of variation of area, pleomorphism index, coefficient of variation of eccentricity and coefficient of variation of circularity between molecular subtypes with Holm correction. Dunn’s post hoc tests were performed following significant Kruskal–Wallis results to compare between all pairs of molecular subtypes. Only significant results are listed. Raw p values and Holm-adjusted p values for multiple comparisons are reported. Scientific notation (E) indicates ×10^power.

| **Dunn-Pairwise Test with Holm Correction – Nuclear Morphometrics** | | | | |
| --- | --- | --- | --- | --- |
| **Metric** | **Group 1** | **Group 2** | **p Value (Raw)** | **p Value (Holm)** |
| Mean nuclear area | P53ABN | NSMP | 6.38E-13 | 2.55E-12 |
| Mean nuclear area | P53ABN | DMMR | 2.14E-23 | 1.28E-22 |
| Mean nuclear area | P53ABN | POLE | 1.80E-20 | 8.98E-20 |
| Mean nuclear area | NSMP | DMMR | 4.01E-05 | 1.20E-04 |
| Mean nuclear area | NSMP | POLE | 6.39E-04 | 1.28E-03 |
| Coefficient of variation of area | P53ABN | NSMP | 1.28E-05 | 7.70E-05 |
| Coefficient of variation of area | P53ABN | DMMR | 9.73E-04 | 3.89E-03 |
| Coefficient of variation of area | NSMP | POLE | 4.71E-05 | 2.36E-04 |
| Coefficient of variation of area | DMMR | POLE | 3.00E-03 | 9.01E-03 |
| Pleomorphism index (PI) | P53ABN | NSMP | 2.60E-03 | 1.30E-02 |
| Pleomorphism index (PI) | P53ABN | DMMR | 1.11E-04 | 6.67E-04 |
| Pleomorphism index (PI) | P53ABN | POLE | 4.31E-03 | 1.72E-02 |
| Coefficient of variation of eccentricity | NSMP | DMMR | 3.36E-03 | 2.01E-02 |
| Coefficient of variation of circularity | P53ABN | NSMP | 5.50E-04 | 1.65E-03 |
| Coefficient of variation of circularity | P53ABN | DMMR | 5.86E-03 | 1.17E-02 |
| Coefficient of variation of circularity | P53ABN | POLE | 4.62E-06 | 1.85E-05 |
| Coefficient of variation of circularity | NSMP | POLE | 7.20E-11 | 4.32E-10 |
| Coefficient of variation of circularity | DMMR | POLE | 2.04E-09 | 1.02E-08 |

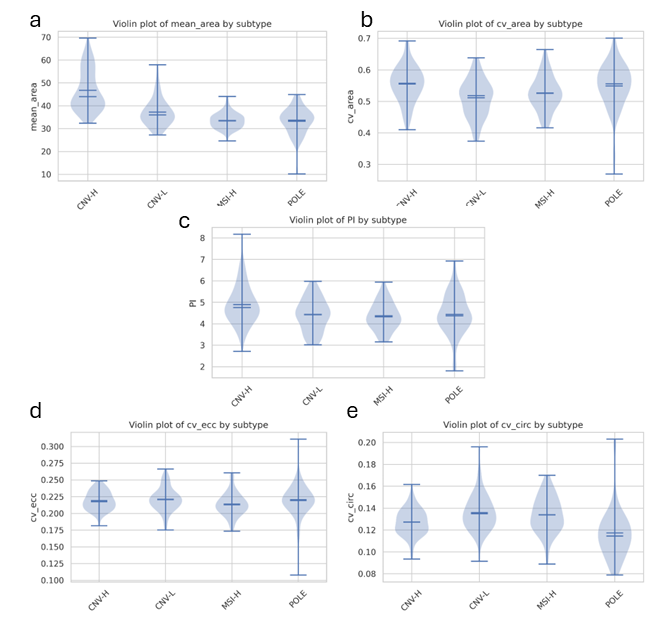

Figure S1: Violin plots of nuclear morphometrics across subtypes, (A) Mean area, (B) coefficient of variation of area, (C) pleomorphism index, (D) coefficient of variation of eccentricity and (E) coefficient of variation of circularity. Subtypes are CNV-H (p53abn), CNV-L (NSMP), MSI-H (dMMR) and POLE.

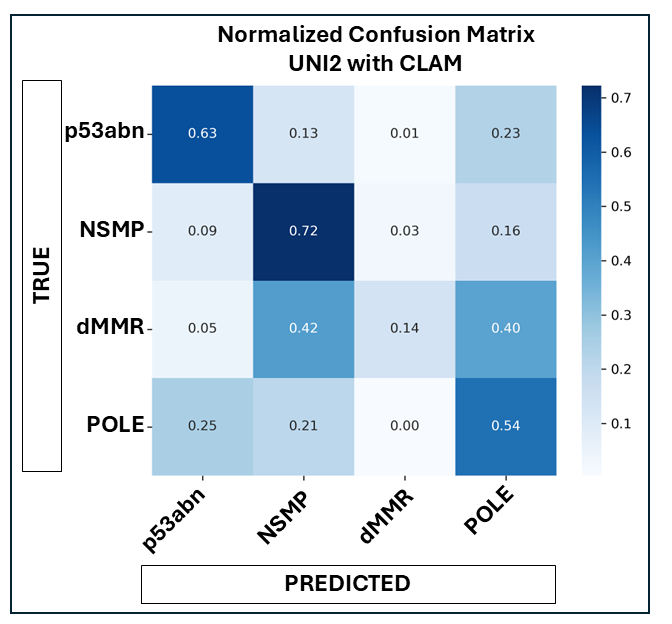

Figure S2: Normalized confusion matrix for UNI2 with CLAM: Confusion matrix showing classification performance of the UNI2 vision transformer with CLAM aggregation for predicting molecular subtypes. Values represent row-normalized proportions of true subtype cases (rows) predicted as each subtype (columns). Darker colors indicate higher proportions.
