## Supplementary Appendix 2 for "Real-World Benchmarking and Validation of Foundation Model Transformers for Endometrial Cancer Subtyping from Histopathology"

**Supplemental Appendix 2**

Table 1 (Page 1): Training configuration and hyperparameter details for the CNN pipeline, transformer pipeline with TransMIL and CLAM.

Page 2: Interpretability analysis, details of settings and methods used.

| TRaining configuration and hyperparameters | | | |
| --- | --- | --- | --- |
| Parameter | **CNN Pipeline** | **Transformer + TransMIL** | **Transformer + CLAM-MB** |
| Input | 512×512 px tiles | 512×512 px tiles → embeddings | 512×512 px tiles → embeddings |
| Freeze ratio | 0.5 | Frozen encoder | Frozen encoder |
| Optimizer | Adam,  LR = 2×10⁻⁴,  WD = 1×10⁻⁵ | AdamW  LR = 1×10⁻⁴,  WD = 1×10⁻² | AdamW,  LR = 1×10⁻⁴,  WD = 1×10⁻² |
| Bag size | N/A | 512 embeddings | 512 embeddings |
| Instance supervision | N/A | None | Yes (K = 32) |
| Dropout | N/A | 0.25 | 0.25 |
| Epochs (max) | 20 | 20 | 20 |
| Early stopping | Patience = 3 (min 5 epochs) | Patience = 5 | Patience = 5 |
| Tile threshold per slide | ≥ 300 tiles | ≥ 300 tiles | ≥ 300 tiles |
| Tile subsampling | ≤ 4,000 tiles | N/A | N/A |
| Aggregation | Mean pooling | Transformer self-attention | Attention pooling + gated classifier |
| Loss function | Cross Entropy | Cross Entropy | Cross Entropy |
| Cross-validation | 5-fold, slide-level stratified | 5-fold, slide-level stratified | 5-fold, slide-level stratified |
| Hardware | 4× NVIDIA RTX A6000 (48 GB) | 4× NVIDIA RTX A6000 (48 GB) | 4× NVIDIA RTX A6000 (48 GB) |

All analyses were implemented in Python 3.11 (SciPy 1.13, Statsmodels 0.15).

**Interpretability Analysis – Settings and Methods**

**Tile Selection**

- Selected 10 highest-predicted WSIs per molecular subtype (based on model probabilities).
- For each WSI, extracted the 8 highest-scoring tiles from attention heatmaps for the correctly predicted subtype.
- Total: 80 tiles per subtype.
- All tiles analyzed at native resolution.

**Segmentation & Classification**

- Performed nucleus-instance segmentation using **HoVer-Net** (Graham et al., 2019) via **TIAToolbox** (Pocock et al., 2022).
- Applied **PanNuke** (Gamper et al., 2019) label schema:
  1. Background
  2. Neoplastic epithelial
  3. Inflammatory
  4. Connective tissue
  5. Dead cells
  6. Non-neoplastic epithelial

**Feature Extraction**

- Restricted quantitative analysis to **neoplastic epithelial nuclei**.
- Computed shape features directly from instance contours:
  - Area (µm²)
  - Perimeter (µm)
  - Eccentricity (from fitted ellipse)
  - Circularity = 4π x Area/(Perimeter^2^)
- Derived summary metrics for each tile:
  - Mean nuclear area (µm²)
  - Size dispersion, coefficient of variation of area (cv_area) = SD(area) / mean(area)
  - Pleomorphism Index (PI) = P90(area) / P10(area)
  - Coefficient of variation of eccentricity (cv_ecc)
  - Coefficient of variation of circularity (cv_circ)

**Aggregation**

- Summarized tile-level indices per slide and per molecular subtype.

**Statistical Analysis**

- Omnibus testing with **Kruskal–Wallis** for differences among four independent subtypes.
- Reported **epsilon-squared effect size**.
- If omnibus test was significant at α = 0.05, performed **Dunn’s post-hoc tests** with Holm correction for multiple comparisons.

**Visualization**

- Cell-type composition shown as **split violin plots** with jittered dot overlays for per-tile proportions.
- Nuclear morphometric distributions displayed as **violin plots**.
